# Protocol for the systematic review of the relationship between Exercise, Gut Microbiota, and serotonin metabolism

**DOI:** 10.1101/2025.05.19.25327922

**Authors:** Yenifer Olivo Martínez, Ángel Antonio Lozano Ariza, Luis Ángel Cardozo Pacheco, Mauricio Ernesto Orozco-Ugarriza

**Affiliations:** Grupo de investigación en Microbiología y Ambiente (GIMA). Universidad de San Buenaventura, Cartagena, Colombia; Grupo Interdisciplinario de Investigación en Educación y Pedagogía (GIEP). Universidad de San Buenaventura, Cartagena, Colombia; Grupo de investigación traslacional en Biomedicina y Biotecnología (GITBB). Corporación para el desarrollo de la Investigación en Biomedicina & Biotecnología, Cartagena, Colombia

**Keywords:** Physical activity, Exercise, Gut microbiota, Gut-brain axis, Serotonin metabolism, Inflammation

## Abstract

**Introduction:** Regular exercise or physical activity can significantly modulate interactions in the gut-brain axis. Currently, it is hypothesized that this may influence the regulation of the serotonergic system and increase the synthesis and bioavailability of serotonin. However, this relationship, as well as the possible mechanisms, remains insufficiently explored.

**Objective:** This review aims to synthesize current evidence on the relationship between physical activity and exercise and their influence on gut microbiota composition and serotonin metabolism, and to analyze how these interactions may impact metabolic and psychological health.

**Methods:** The protocol is prepared following the preferred reporting items for systematic reviews and meta-analyses (PRISMA-P) guidelines. We will perform a systematic review of literature published between January 2016 and July 2024, using the databases PubMed, Google Scholar. In addition, manual searches will be carried out through related articles, and references to included articles. The main findings, including primary and secondary outcome measures, will be extracted from all eligible studies using a standardized instrument. Two authors will independently screen titles and abstracts, review full texts, and collect data. Disagreements will be resolved by discussion, and a third reviewer will decide if there is no consensus. We will synthesize the results using a narrative or a meta-analytic approach, depending on the heterogeneity of the studies.

**Expected results:** It is expected that this systematic review will provide a comprehensive and up-to-date overview of the current understanding of the relationship between exercise, gut microbiota, and serotonin metabolism. Furthermore, the study will identify potential gaps in knowledge that warrant further investigation through future research.

**Conclusions:** Here, we present the protocol for a systematic review that will consider all existing evidence from peer-reviewed sources relevant to the primary and secondary outcomes related to the relationship between exercise, gut microbiota, and serotonin metabolism.

**Systematic review registry:** This study was registered at the International Prospective Registry for Systematic Reviews (PROSPERO registration number: CRD42024606591).

## Introduction

Multiple studies have established the beneficial effect of physical exercise or physical activity on a wide range of biological functions in the host. Research has shown that exercise or physical activity can significantly modulate gut-brain axis interactions. Specifically, scientific evidence has demonstrated that physical activity can shift microbial profiles, enhancing diversity and functional pathways that may affect neurotransmitter production, among which serotonin is crucial for mood regulation, gastrointestinal function, and metabolic homeostasis. [1,2]. These interactions are particularly relevant given serotonin’s dual role as a neurochemical and a peripheral signaling molecule, with approximately 90% of serotonin synthesized in the gut. However, beyond those functions, we noted that the relationship between exercise-induced serotonin effects and gut microbiota composition, as well as the possible mechanisms involved by which it exerts its effect through gut-brain interactions, remains insufficiently explored.

Exercise-induced changes in gut microbiota and serotonin levels have implications for a wide range of conditions, including inflammatory disorders, metabolic diseases, and psychological conditions such as anxiety and depression [3,4]. However, despite its potential, the underlying mechanisms linking exercise to microbiota-mediated serotonin production remain poorly understood. Variability in study populations, exercise protocols, and methods for measuring microbiota and serotonin complicates efforts to synthesize evidence and draw definitive conclusions.

Furthermore, while studies have demonstrated that exercise can stimulate serotonin synthesis by influencing gut microbial pathways, inconsistencies in findings raise questions about the specific taxa and microbial metabolites involved, as well as the extent to which these effects translate into measurable health benefits [5,6]. This knowledge gap underscores the need for a comprehensive review of the existing literature to elucidate how physical activity modulates gut microbiota and serotonin levels and to explore its broader implications for health and disease prevention.

This systematic review protocol presents a detailed plan to synthesize the evidence on this topic. By examining the interplay between exercise, gut microbiota, and serotonin, this review will focus on studies that explore these interactions and aim to synthesize current evidence on the influence of different types, intensities, and durations of physical activity or exercise on variations in gut microbiota composition and serotonin metabolism. The findings expected that this systematic review would provide a comprehensive and up-to-date overview of the state-of-the-art relationship between physical activity or exercise and its influence on gut microbiota composition and serotonin levels in adults.

## Methods

### Design and registration

This systematic review protocol has been conducted following the recommendations of the Preferred Reporting Items for Systematic Reviews and Meta-Analysis (PRISMA-P) guidelines [7]. This review protocol was registered in the Prospective Register of Systematic Reviews platform (PROSPERO) [8], (registration number CRD42024606591), and can be accessed at https://www.crd.york.ac.uk/PROSPERO/view/CRD42024606591.

See (S1 Checklist) for the completed PRISMA-P checklist. We will only make changes to the study protocol if necessary and report them on the PROSPERO record.

### Search strategy

The search strategy will be conducted comprehensively across electronic databases, including PubMed and Google Scholar. Controlled terms (i.e. MeSH) and relevant keywords will be used to identify relevant studies related to Exercise, Gut Microbiota, and Serotonin Metabolism. To expand and refine the search, we will use Boolean operators (such as AND, OR, NOT) and truncations, as per the specificities of each database. The search strategy will be constructed using combinations of the following terms: “Exercise” OR “Physical exercise” OR “Physical Activity” AND “Gut Microbiome” OR “Gut Microbiota” OR “gut Microbes” OR “Intestinal Microbiome” OR “Intestinal Microbiota” OR “Gut Health” AND “Serotonin” OR “5-hydroxytryptamine” OR “Sertoninergic system”

In addition, the references of all relevant primary articles will be reviewed to include studies based on content relevance without any bias toward the author or journal and any articles that may have been missed in electronic searches.

The search will be limited to studies published between 2016 and 2024, aiming to obtain the most up-to-date evidence on the topic of interest. Only studies published in English or Spanish will be included. However, efforts will be made to identify and access relevant studies published in other languages if possible. The details of the search strategy, including the terms used and filters applied, will be recorded to ensure transparency and reproducibility of the systematic review.

The pilot search will be adapted to each database, avoiding any inconsistencies that may affect data extraction and ensuring that the final search strategy is correctly adapted. The details of the search strategy, including the terms used and filters applied, will be recorded to ensure transparency and reproducibility of the systematic review.

### Eligibility criteria

We will include studies that meet the following criteria:

#### Population

This review will include studies involving both healthy adults and those with specific health conditions. This approach enables the exploration of how exercise and physical activity impact individuals differently based on their health status, specifically on gut microbiota and serotonin levels.

#### Intervention(s) or exposure(s)

The interventions and exposures for this systematic review will include studies that prescribe both structured exercise and general physical activity, encompassing modalities such as supervised or individualized exercise, resistance training, respiratory muscle training, walking, or a combination of these modalities.

#### Comparator(s) or control(s)

This will be eligible for inclusion, studies that examine different types of physical activity or studies with control group in which exercise or physical activity is not intervention or exposure will be eligible for inclusion.

#### Outcome

Any outcome related to changes in gut microbiota composition and serotonin levels in response to exercise or physical activity interventions in adults.

### Main outcomes

The primary outcomes of this systematic review are changes in gut microbiota composition and serotonin levels in response to exercise or physical activity interventions in adults.

#### Gut Microbiota Composition

This outcome focuses on shifts in the diversity, abundance, and specific bacterial taxa within the gut microbiota as measured by different microbiota profiling techniques. Standard metrics include microbial alpha and beta diversity indices, the presence of beneficial bacterial species, and changes in microbial metabolic functions.

#### Serotonin levels

This outcome evaluates the quantitative concentration of serotonin (5-hydroxytryptamine or 5-HT) in body fluids, principally blood, such as an indicator of changes in response to exercise or physical activity interventions.

##### Measures of effect

The measures of effect for this systematic review will focus on quantifying the impact of exercise or physical activity on gut microbiota composition and serotonin levels. The measures of effect will include:

#### Diversity and Relative Abundance of Gut Microbiota

Gut microbiota composition will be measured by quantifying alpha diversity (Shannon, Simpson) and beta diversity (Bray-Curtis dissimilarity), as well as estimating the relative abundance of microbial lineages at multiple phylogenetic resolutions (i.e., phylum, genus and species-like resolution) and expressed as percentages or ratios, accounting for evolutionary phylogenetic timescales, if possible.

#### Serotonin levels

Measures of effect will be based on available literature and include summary statistics to capture changes in serotonin concentration levels before and after exercise or physical activity interventions. The specific measures of effect may vary according to the diagnostic approaches and laboratory methods used in each study.

### Additional outcomes

In addition to serotonin levels measured in blood, this systematic review will include studies that explore other additional outcomes related to serotonin metabolism, which may include but are not limited to, the following data: levels of serotonin in other biological samples; levels of serotonin metabolites; levels of expression of genes or proteins related to the serotonergic system.

#### Measures of effect

The specific measures of effect may differ according to the literature data nature of each study.

### Study design

The types of studies eligible for inclusion in this systematic review are original articles that declare randomized controlled trials (RCTs), non-randomized controlled trials design of study, as well as analytical observational studies (cohort, case-control, and cross-sectional studies) that explore the relationship between exercise or physical activity and changes in gut microbiota composition and serotonin levels. We will exclude case reports, case series, letters, editorials, and reviews that do not provide original data.

### Publication date

The search will be limited to studies published between January 2016 and December 2024. To ensure the inclusion of the most up-to-date evidence on the relationship between exercise, gut microbiota, and serotonin metabolism, the search may be extended if necessary. This will include manual searches database, reviewing the bibliographic references of included studies, and considering expert recommendations.

### Language

We will include studies published in English or Spanish, as these are the languages that the reviewers can read and understand. However, efforts will be made to identify and access relevant studies published in other languages if possible.

### Literature screening and study selection

The screening and study selection process will follow a structured 3-step methodology (titles, abstracts, and full texts) to determine the eligibility of studies based on predefined inclusion and exclusion criteria. First, duplicates will be identified and removed from the retrieved search results using Mendeley desktop software. Second, two reviewers (MEOU and YOM) will independently screen the titles and abstracts of all identified studies. Finally, the articles passing the initial screening stage will proceed to detail a full-text review. Any disagreement between the reviewers will be resolved by discussion or, if needed, by consulting a third reviewer (ALA), who will decide if there is no consensus.

We plan to make use of a PRISMA-compliant flowchart to summarize the article screening process, detailing the number of studies included and excluded at each stage, along with the reasons for exclusion of studies after the full-text review, as depicted in the S2 File. This process will follow the PRISMA guidelines for reporting systematic reviews [9].

### Data extraction

Data will be collected independently by three individual reviewers from each eligible publication, and the extracted data will be recorded using a standardized data extraction form or template developed in Microsoft Excel (See S3 File). Efforts will be made to contact study investigators to request unreported data or additional details for missing data.

The implementation of the data extraction form will be carried out in four stages: 1) development of a data extraction form, 2) peer-validation of the data extraction form, 3) pilot test of the data extraction form, and 4) final data extraction.

### Data items

In the systematic review, the following study characteristics or data will be extracted from the included studies, which may cover, but are not limited to, the following categories:

#### Identification of the study

Information related to the identification of the study on the first author and year of publication, article title, short citation, and country.

#### Methods

This will include the study objectives, study design, sample size, population characteristics (mean age, sex, health status, inclusion/exclusion criteria), intervention specifics (type, duration, frequency, and intensity of exercise or physical activity), diagnostic methods and a summary of the PICOS (Population, Intervention, Comparison, Outcome [10], if applicable.

#### Main findings

This will include primary and secondary outcome measures (such as gut microbiota composition and serotonin levels), results description (summary statistics and subgroup analyses), and a summary of key findings, a One-sentence conclusion of the study, and any limitations noted in the study.

### Methodological quality assessment

For the systematic review, the method of assessing the risk of bias will involve the following steps:

#### Independent assessment

Two reviewers will independently evaluate the methodological quality of the included studies.

#### Disagreement resolution

Any disagreements between reviewer’s assessments will be resolved through discussion and consensus. In cases where consensus cannot be reached, a third reviewer will be involved in reaching a final agreement.

#### Study selection and data extraction

Two reviewers will independently assess the search results and select high-quality literature for inclusion in the review. Multiple reviewers will be involved in the data extraction process to minimize the risk of errors.

#### Blinding

The study selection, data extraction, and risk of bias assessment will be performed without blinding the reviewers to the study authors or the journal of publication.

#### Quality and Bias Assessment Tools

If possible, the included studies will be systematically evaluated using tools for assessing study designs, focusing on selection methods, comparability, and outcome assessment.

These measures are implemented to ensure the rigor and reliability of the review process. By involving multiple independent reviewers and resolving disagreements through discussion and consensus, the risk of bias in the included studies will be assessed transparently and comprehensively.

### Strategy for data synthesis

We planned to synthesize the data using a systematic narrative synthesis approach to summarize the key findings from the included studies, along with population characteristics, interventions, outcomes of interest, methodological quality, and any identified limitations. This synthesis will offer a comprehensive understanding of the topic that integrates the findings and characteristics of the studies.

To facilitate analysis and interpretation, we will categorize these findings into relevant subgroups or subsets grouped into three categories based on their primary focus: (1) the effects of physical activity or exercise on gut microbiota composition; (2) the influence of physical activity or exercise on serotonin levels or metabolism; and (3) studies examining the triadic interaction between exercise, gut microbiota, and serotonin. This grouping will allow for a more structured synthesis of the evidence and help identify patterns, divergences, and gaps across different research approaches.

Additionally, we will employ descriptive statistics to analyze study characteristics and outcomes. We will not be employing meta-regression, tests of interaction, or statistical modeling, as the aim of this review is to provide a qualitative synthesis, focusing on summarizing and interpreting the available evidence.

### Ethics

This is a systematic review that will use published data and does not require ethical approval.

### Status of the study and dissemination plan

This systematic review is currently in the pilot and preliminary search phase, pending approval of the Registered Report Protocol. We aim to complete the project, publish the results in a peer-reviewed journal, and report the findings within the next 12 months. The final report will follow the updated PRISMA guidelines and be registered on the PROSPERO website. All the data will be accessible as supplementary information after the review. In addition, we plan to present the main findings at academic conferences and relevant professional meetings focused on exercise science, gut microbiota, and neurobiology.

## Discussion

The relationship between physical activity, gut microbiota composition, and serotonin metabolism represents a growing area of interest in understanding how lifestyle factors influence mental and physical health. Exercise is known to promote microbial diversity and enhance the production of beneficial microbial metabolites, potentially supporting the synthesis of serotonin, a neurotransmitter involved in mood regulation, immune function, and metabolic processes [1,11]. Recent research highlights that exercise-induced changes in the gut microbiota may contribute to reducing systemic inflammation and improving neurochemical balance, offering potential non-pharmacological pathways for managing psychological and metabolic conditions. However, inconsistencies in study findings, driven by variability in exercise types, intensity, and study populations, complicate understanding these interactions [12,13].

Despite advances in microbiota research and its integration into exercise science, there is limited consensus on the mechanisms linking gut microbial shifts to serotonin metabolism. Differences in study methodologies, such as variations in microbiota profiling techniques and serotonin measurement methods, pose challenges in synthesizing evidence. Furthermore, while some studies suggest exercise enhances serotonin synthesis through microbial pathways, others report conflicting results, particularly regarding the specific microbial taxa and metabolites involved. These uncertainties hinder the development of targeted interventions and highlight the need for a comprehensive review of the current evidence [5,6].

This systematic review protocol aims to address these gaps by systematically evaluating the effects of physical activity on gut microbiota composition and serotonin metabolism. This review will provide unbiased evidence on how exercise modulates these interrelated systems by detailing the protocol development, search strategy, study selection, data extraction, and quality assessment. The knowledge obtained from this systematic review can contribute to the application of non-pharmacological approaches and the development of personalized medicine strategies to manage conditions associated with serotonin dysregulation. On the one hand, this systematic review will provide a comprehensive and updated overview of the state-of-the-art knowledge of how physical activity or exercise influences the gut microbiota composition and serotonin levels in this population.

Finally, we hope to identify existing knowledge gaps that may form the basis of possible future lines of research on various aspects of the relationship between physical activity or exercise and its influence on gut microbiota composition and serotonin levels in adults, that may contribute to understanding of their combined influence on metabolic, inflammatory, and psychological health.

## Conclusion

The beneficial effect of physical exercise or physical activity on a wide range of biological functions in the host has been well established. Currently, research has shown that exercise or physical activity can significantly modulate gut-brain axis interactions. Specifically, it has been demonstrated that physical activity can shift microbial profiles, enhancing diversity and functional pathways that may affect neurotransmitter production. However, beyond those functions, we noted that the relationship between exercise-induced serotonin effects and gut microbiota composition, as well as the possible mechanisms involved by which it exerts its effect through gut-brain interactions, remains insufficiently explored.

Therefore, it is essential to conduct a systematic review that synthesizes the available evidence on the primary and secondary outcomes related to with diversity and relative abundance of gut microbiota composition, serotonin metabolism (levels of serotonin, levels of serotonin metabolites, and levels of expression of genes or proteins related to the serotonergic system), intervention specifics (type, duration, frequency, and intensity of exercise or physical activity).

In this protocol, we describe the methods and criteria we will use to perform a comprehensive and up-to-date search, selection, extraction, quality assessment, and synthesis of data from peer-reviewed publications relevant to the primary and secondary outcomes of interest. We expect that this systematic review will provide a state-of-the-art overview of the current knowledge of how physical activity or exercise influences gut microbiota composition and serotonin levels. It will also give a clearer understanding of their combined influence on metabolic, inflammatory, and psychological health, identify the existing knowledge gaps, and suggest possible future lines of research.

## Supporting information

S1 Checklist. Preferred Reporting Items for Systematic Review and Meta-Analysis Protocols (PRISMA-P checklist)

S2 File. PRISMA 2009 flow diagram

S3 File. Data extraction form for systematic review

S4 File. PROSPERO registration

## Data Availability

All data from this study will be made available in the online databases upon study completion.

## Supporting information

S2 File. PRISMA 2009 flow diagram.

S3 File. Data extraction form for systematic review. S4 File. PROSPERO registration.

## Acknowledgments

The authors would like to thank all members of the group for their support and help.

## Author contributions

**Mauricio Ernesto Orozco-Ugarriza:** Contributed equally to this work with Yenifer Olivo-Martínez. Roles: Conceptualization, Data curation, Methodology, Supervision, Validation, Visualization, Writing – original draft, Writing – review & editing.

**Yenifer Olivo-Martínez:** Contributed equally to this work with: Mauricio Ernesto Orozco-Ugarriza. Roles: Conceptualization, Data curation, Validation, Visualization, Writing – original draft, Writing – review & editing.

**Ángel Antonio Lozano Ariza:** Roles: Writing – review & editing.

**Luis Ángel Cardozo Pacheco:** Roles: Writing – review & editing.

## Availability of data and materials

All data from this study will be made available in the online databases upon study completion.

## Ethics approval and consent to participate

This study has no ethics approval needed.

## Consent for publication

No need for consent for research and publication.

