## Supplementary material for "Protocol for the systematic review of the relationship between Exercise, Gut Microbiota, and serotonin metabolism": S3 File. Data extraction form for systematic review

**S3 Table: Data Extraction Form for Systematic Review on the Relationship Between Exercise, Gut Microbiota, and Serotonin Metabolism.**

| **General Information:** | |
| --- | --- |
| **DATA COLLECTION ITEM** | **ITEM DESCRIPTION / SUB-GROUPS** |
| COUNTRY | Country where the study was conducted |
| TYPE OF STUDY | Type of research design (e.g., randomized controlled trial, longitudinal study, cross-sectional study) |
| TIME PERIOD AND SETTING OF THE STUDY | Duration and context in which the study took place (e.g., lab-based, clinical, community) |

**Methods**

| **DATA COLLECTION**  **ITEM** | **ITEM DESCRIPTION / SUB-GROUPS** |
| --- | --- |
| **INTERVENTION** | Type of physical activity or exercise (e.g., aerobic, resistance, endurance, HIIT) |
| **DESCRIPTION OF INTERVENTION** | Detailed description of the exercise protocol (frequency, intensity, duration, type) |
| **THE SETTING OF THE INTERVENTION** | Context of intervention (e.g., clinical setting, laboratory, at-home, supervised) |
| **AIM / OBJECTIVES** | Study objectives related to the impact of exercise on gut microbiota, serotonin levels, or their interaction |
| **PARTICIPANT CHARACTERISTICS** | - Participant demographics (age, sex, health status) - Comparator details: Description of control or alternative intervention group - Inclusion Criteria: Eligibility for study participation - Exclusion Criteria: Criteria for non-inclusion |
| **OUTCOME MEASURES** | Outcomes related to: - Gut microbiota composition (e.g., diversity indices, abundance of specific taxa) - Serotonin levels or metabolism (e.g., blood, plasma, or CNS levels; expression of serotonergic genes) - Interaction outcomes (e.g., changes mediated by gut-brain axis) |
| **SOURCE OF DATA** | Biological sample source (e.g., stool, blood, CNS tissue); questionnaire or instrument used |
| **DATA ANALYSIS** | Statistical methods used (e.g., microbiome sequencing analysis, metabolomics, correlation/regression analysis) |

**Results**

| **DATA COLLECTION ITEM** | **ITEM DESCRIPTION / SUB-GROUPS** |
| --- | --- |
| **STUDY PARAMETERS** | Key variables measured and reported related to exercise, gut microbiota, and serotonin |
| **OUTCOMES** | - Mean values or changes in outcomes - Differences between groups (e.g., intervention vs. control) |
| **CHARACTERIZING HETEROGENEITY** | Variability in study outcomes (e.g., population characteristics, exercise protocols, microbiome measurement techniques) |

**Discussion**

| **DATA COLLECTION ITEM** | **ITEM DESCRIPTION / SUB-GROUPS** |
| --- | --- |
| **STUDY FINDINGS** | **Summary of major findings related to the interaction between exercise, gut microbiota, and serotonin** |
| **LIMITATIONS** | **Reported limitations (e.g., sample size, study duration, lack of microbiota analysis depth)** |
| **GENERALISABILITY** | **Extent to which findings can be applied to other populations or contexts** |
| **CURRENT KNOWLEDGE** | **Alignment with or contrast to existing literature on the gut-brain axis, exercise physiology, or serotonin regulation** |
| **IMPLICATIONS FOR FUTURE RESEARCH OR PRACTICE** | **Suggestions related to future directions, clinical application, or development of non-pharmacological interventions** |
| **CONCLUSION** | **Summary conclusion of the study’s contribution to the topic** |
