## Supplementary material for "Protocol for the systematic review of the relationship between Exercise, Gut Microbiota, and serotonin metabolism": S4 File. PROSPERO registration

NIHR | National Institute for Health and Care Research

PROSPERO
International prospective register of systematic reviews

### Systematic review of the relationship between Exercise, Gut Microbiota, and Serotonin Metabolism.

*Mauricio Ernesto Orozco Ugarriza, Yenifer Olivo Martínez, Angel Antonio Lozano Ariza*

To enable PROSPERO to focus on COVID-19 submissions, this registration record has undergone basic automated checks for eligibility and is published exactly as submitted. PROSPERO has never provided peer review, and usual checking by the PROSPERO team does not endorse content. Therefore, automatically published records should be treated as any other PROSPERO registration. Further detail is provided here.

**Citation**
Mauricio Ernesto Orozco Ugarriza, Yenifer Olivo Martínez, Angel Antonio Lozano Ariza. Systematic review of the relationship between Exercise, Gut Microbiota, and Serotonin Metabolism.. PROSPERO 2024 Available from https://www.crd.york.ac.uk/PROSPERO/view/CRD42024606591

#### REVIEW TITLE AND BASIC DETAILS

##### Review title
Systematic review of the relationship between Exercise, Gut Microbiota, and Serotonin Metabolism.

##### Review objectives
In adults (healthy or with disease) (P), how does regular exercise or physical activity (I) compared to a sedentary lifestyle (C) affect gut microbiota and serotonin levels (O)?
P (Population): Any adult (healthy or with conditions disease)
I (Intervention): Regular exercise or physical activity (any form of structured exercise)
C (Comparator): Sedentary lifestyle or no exercise intervention
O (Outcome): Changes in gut microbiota and serotonin levels
The gut microbiota is a complex ecosystem with significant implications for host digestion, metabolism, immune function, the maintenance of intestinal health, and overall health. Shifts in microbiota composition, abundance, and diversity have been associated with different metabolic outcomes. Currently, research has shown that the gut microbiota influences the expression of

serotonergic genes in the host, thereby affecting serotonin's bioavailability and its functions. Moreover, regular exercise is hypothesized to influence gut microbiota through various mechanisms, including the modulation of inflammatory markers, production of short-chain fatty acids, and improvement in metabolic biomarkers such as serotonin. This review will focus on studies that explore these interactions to determine whether different types, intensities, and durations of physical activity correlate with variations in the composition of gut microbiota and serotonin metabolism.

#### ELIGIBILITY CRITERIA

##### Condition or domain being studied

This systematic review focuses on the relationship between physical activity or exercise and its influence on gut microbiota composition and serotonin levels in adults, emphasizing how these factors may interact to affect metabolic and psychological health. Key areas of interest include exploring physical activity or exercise's role in modifying microbiota composition and serotonin regulation.

Relevance of the Study Domain: Currently, research has shown that exercise or physical activity can significantly modular the gut-brain axis interactions. Exercise can modify microbiota composition, and influence the metabolites produced by gut bacteria, potentially contributing to reversing conditions associated with obesity, metabolic diseases, poor diets, as well as certain neurological and behavioral disorders. Moreover, physical activity—particularly aerobic exercise—may influence regulating the serotonergic system and increasing the synthesis and availability of serotonin.

Understanding the interaction of this triadic relationship between exercise, gut microbiota, and serotonin, is particularly significant for the application of non-pharmacological approaches and contributes to the development of strategies of personalized medicine. This knowledge can aid in managing a wide range of conditions associated with serotonin dysregulation, including inflammation, metabolic diseases, and psychological disorders.

Data will be collected independently by two reviewers (YOM and MEOU) from each eligible publication, and the extracted data will be recorded using a standardized data extraction form or template developed in Microsoft Excel. Efforts will be made to contact study investigators to request unreported data or additional details for missing data.

##### Risk of bias (quality) assessment

For the systematic review, the method of assessing the risk of bias will involve the following steps:
Independent assessment: Two reviewers will independently evaluate the methodological quality of the included studies.
Disagreement resolution: Any disagreements between reviewer's assessments will be resolved through discussion and consensus. In cases where consensus cannot be reached, a third reviewer will be involved to reach a final agreement.
Study selection and data extraction: Two reviewers will independently assess the search results and select high-quality literature for inclusion in the review. Multiple reviewers will be involved in the data extraction process to minimize the risk of errors.
Blinding: The study selection, data extraction, and risk of bias assessment will be performed without blinding of the reviewers to the study authors or the journal of publication.
Quality and Bias Assessment Tools: If possible, the included studies will be systematically evaluated using tools for assessing study designs, focusing on selection methods, comparability, and outcome assessment.
These measures are implemented to ensure the rigor and reliability of the review process. By involving multiple independent reviewers and resolving disagreements through discussion and consensus, the risk of bias in the included studies will be assessed transparently and comprehensively.

#### PLANNED DATA SYNTHESIS

##### Strategy for data synthesis

We planned to synthesize the data using a systematic narrative synthesis approach, which will summarize the primary findings from the included studies, as well as the population characteristics interventions, outcomes of interest, and methodological quality of the studies and any identified limitations.
Overall, the narrative synthesis will provide a comprehensive understanding of the topic by presenting a cohesive narrative that integrates the findings and characteristics of the included studies.
We will not be employing meta-regression, tests of interaction, or statistical modeling since the objective is to provide a qualitative synthesis rather than a meta-analysis.

##### Analysis of subgroups or subsets

If possible, we will categorize these findings into relevant subgroups or subsets depending on data availability to facilitate a clearer presentation of the synthesized data. Additionally, we will employ descriptive statistics to analyze the results, where feasible.

#### REVIEW AFFILIATION, FUNDING AND PEER REVIEW

##### Review team members


- Professor Mauricio Ernesto Orozco Ugarriza, Universidad de San Buenaventura Cartagena
- Dr Yenifer Olivo Martínez, Universidad de San Buenaventura Cartagena
- Professor Angel Antonio Lozano Ariza, Universidad de San Buenaventura Cartagena


##### Review affiliation

Universidad de San Buenaventura Cartagena,

##### Funding source

**None<\/b>**

##### Named contact

Mauricio Ernesto Orozco Ugarriza. Colombia. Dg. 32 #No. 30-966, Ternera, Cartagena de Indias. CP: 130010

#### TIMELINE OF THE REVIEW

##### Review timeline

Start date: 03 February 2025. End date: 30 June 2025

##### Date of first submission to PROSPERO

01 November 2024

##### Date of registration in PROSPERO

14 November 2024

#### CURRENT REVIEW STAGE

##### Publication of review results

The intention is not to publish the review once completed.

##### Stage of the review at this submission

| Review stage | Started | Completed |
|---|---|---|
| Pilot work | | |
| Formal searching/study identification | | |
| Screening search results against inclusion criteria | | |
| Data extraction or receipt of IP | | |
| Risk of bias/quality assessment | | |
| Data synthesis | | |

This systematic review, as of now, has not yet commenced. Before initiating the formal search, pilot searches will conducted across various databases, tailoring the strategy to each to prevent inconsistencies that might impact data extraction.

##### Review status

The review is currently planned or ongoing.

#### ADDITIONAL INFORMATION

##### PROSPERO version history

- Version 1.1 published on 14 Nov 2024
- Version 1.0 published on 14 Nov 2024

##### Review conflict of interest

None known

##### Country

Colombia, Spain

##### Medical Subject Headings

Adult; Biological Availability; Biomarkers; Digestion; Exercise; Fatty Acids, Volatile; Gastrointestinal Microbiome; Health Status; Humans; Immunity; Microbiota; Sedentary Behavior; Serotonin

##### Disclaimer

The content of this record displays the information provided by the review team. PROSPERO does not peer review registration records or endorse their content.

PROSPERO accepts and posts the information provided in good faith; responsibility for record content rests with the review team. The owner of this record has affirmed that the information provided is truthful and that they understand that deliberate provision of inaccurate information may be construed as scientific misconduct.

PROSPERO does not accept any liability for the content provided in this record or for its use. Readers use the information provided in this record at their own risk.

Any enquiries about the record should be referred to the named review contact
